# Stage-specific gut microbiome shifts across the Type 2 Diabetes Mellitus spectrum: A systematic review and meta-analysis

**DOI:** 10.64898/2026.01.20.25341999

**Authors:** Sanaa Harrass, Shaimaa Ali, Mariam Elshweikh, Ricardo Franco-Duarte, Thilini N. Jayasinghe

## Abstract

**Aims:** The gut microbiome has been implicated in type 2 diabetes progression, but reproducible biomarkers across studies remain limited due to technical and population heterogeneity. This study investigated whether specific gut microbiome shifts occur progressively across stages of type 2 diabetes.

**Methods:** We systematically reanalysed 16S rRNA datasets from 12 published studies (n=1,247 samples) after quality control, examining five groups (healthy controls, prediabetes (PD), new-onset type 2 diabetes, established type 2 diabetes, and type 2 diabetes with complications. Sequencing reads were quality-filtered, denoised, and resolved into amplicon sequence variants with genus-level taxonomic assignments using the SILVA database. Centered log-ratio (CLR)–transformed abundance data were analysed using PERMANOVA, meta-analysis with leave-one-study-out validation, differential abundance testing (Wilcoxon and ANCOM), and Random Forest classification. Eligible studies were identified through comprehensive searches of PubMed, Ovid Medline and Web of Science from June 2010 – June 2025 using predefined inclusion and exclusion criteria following PRISMA 2020 guidelines. Studies were investigated by two independent reviewers and included if they provided 16S rRNA data on adults across diabetes stages. Study quality was assessed based on metadata completeness and raw data availability. This systematic review and meta-analysis was registered in the Open Science Framework (OSF; registration https://osf.io/eth7a; embargoed until October 2026) and conducted according to PRISMA guidelines.

**Results:** Early disease transitions showed minimal microbiome alterations, with only 4 genera, (notably enrichment of *Allisonella* and *Escherichia–Shigella)* were significantly different between healthy and PD (q < 0.05), and no significant genera between PD and new-onset type 2 diabetes. Advanced disease exhibited robust dysbiosis, with 9 genera differentially abundant in type 2 diabetes vs complicated type 2 diabetes and 5 genera in healthy vs complicated type 2 diabetes comparisons. Complicated type 2 diabetes was characterised by enrichment of *Hungatella* and *[Clostridium] innocuum* group and depletion of *Faecalibacterium* and compared to both uncomplicated type 2 diabetes and healthy controls. Random Forest classification achieved poor performance for early contrasts (AUC ≤ 0.79) but strong discrimination for advanced disease (type 2 diabetes vs complicated type 2 diabetes: AUC = 0.89; Healthy vs complicated type 2 diabetes: AUC = 0.96).

**Conclusion:** Gut microbiome alterations are subtle and inconsistent in early dysglycemia but become pronounced and reproducible with diabetic complications, suggesting microbiome-based biomarkers may be most clinically useful for identifying disease progression rather than early detection. Limitations include heterogeneity of sequencing methods and reliance on 16S rRNA data, which may restrict taxonomic and functional resolution. To our knowledge, this is the first meta-analysis to systematically evaluate gut microbiome alterations across multiple clinical stages of type 2 diabetes progression.

## Introduction

The human gut microbiome plays a central role in maintaining metabolic homeostasis through its complex interactions with host physiology [1]. Gut microbes harvest energy from dietary components, regulate immune function, and produce metabolites that influence systemic inflammation and metabolism [2]. These microbial communities also communicate with distant organs through pathways such as the gut–brain axis, gut–liver, gut–kidney, and gut–heart axes. These pathways link the microbiome to neurological function, bile acid and glucose metabolism, renal health, and cardiovascular risk, underscoring its broad systemic impact [3–6].

Disruptions in gut microbial composition, commonly termed “dysbiosis,” have been implicated in numerous metabolic disorders, including type 2 diabetes mellitus (type 2 diabetes) and its complications [7, 8]. The progression from healthy glucose metabolism through prediabetes to overt diabetes and eventually to complicated diabetes represents a continuum of metabolic dysfunction that may be reflected in corresponding shifts in gut microbial communities [9]. Characterising these changes could provide valuable insights into disease pathogenesis and identify microbial biomarkers for early detection, risk stratification, and monitoring of disease progression [9, 10].

Individual case–control studies investigating the gut microbiome in diabetes have reported various disease-associated microbial alterations [11–13]. However, findings are often inconsistent, partly due to differences in study populations, methodologies, sequencing pipelines, and geographic variation [14]. The concept of “dysbiosis” itself is inconsistently defined across studies, complicating efforts to identify reproducible microbial signatures that clearly distinguish different stages of disease [15].

Meta-analysis provides a powerful approach to address these challenges by synthesising data across multiple cohorts, thereby reducing study-specific biases and identifying consistent, robust microbial associations [16]. Previous meta-analyses in other disease contexts, such as colorectal cancer and inflammatory bowel disease, have successfully revealed universal microbial markers that transcend individual studies [15, 17, 18]. Yet, comprehensive meta-analyses focused specifically on the diabetes spectrum, from healthy individuals through prediabetes to type 2 diabetes and complicated diabetes remain limited.

Therefore, this meta-analysis provides the first complete synthesis of evidence on gut microbiota alterations spanning the full continuum of diabetes progression. In this study, we conducted a comprehensive meta-analysis of published 16S rRNA gene amplicon sequencing datasets to investigate gut microbiome alterations across the diabetes progression spectrum. By reprocessing and analysing multiple independent cohorts through a unified workflow, we aimed to (i) identify consistent microbial signatures associated with healthy, prediabetic, diabetic, and type 2 diabetes with complications (ii) determine whether distinct disease stages are characterised by specific microbial shifts or by non-specific microbial responses to metabolic dysfunction; and (iii) evaluate the potential of gut microbial markers as robust, non-invasive biomarkers for early detection and monitoring of diabetes progression (iv) assess whether genus-level aggregation or ASV-level resolution provides greater sensitivity for detecting stage-specific microbiome alterations.

## Results

To systematically evaluate gut microbiome alterations across the diabetes continuum, we integrated 12 publicly available 16S rRNA sequencing datasets encompassing 1,520 stool samples. Eligible studies were identified through systematic searches of PubMed, Ovid MEDLINE, and Web of Science covering the period from June 2010 to 15^th^ of June 2025. Search terms combined keywords and MeSH headings for “gut microbiome,” “microbiota,” “type 2 diabetes,” “prediabetes,” and “diabetic complications.” Inclusion criteria required human stool 16S rRNA datasets with available raw sequencing data and metadata defining diabetes status; non-human studies or those lacking accessible data were excluded. Two reviewers independently screened titles, abstracts, and datasets, with discrepancies resolved by consensus. The protocol was registered in the Open Science Framework (OSF; registration https://osf.io/eth7a; embargoed until October 2026) and conducted according to PRISMA guidelines. We initially included 16 studies [19–34] then 4 were excluded from the meta-analysis due to quality and comparability issues [19, 24, 30, 31]. The included studies had participants spanning healthy controls, prediabetes (PD), newly diagnosed type 2 diabetes (type 2 diabetes_new), established type 2 diabetes, and type 2 diabetes with complications (type 2 diabetes_comp), where complicated cases were pooled from diabetic nephropathy (DN), diabetic retinopathy (DR), and proliferative diabetic retinopathy (PDR). All datasets represented diverse geographic regions and sequencing platforms but were processed using standardized QIIME2 (v2024.2) pipeline [15]. Downstream analyses based on normalised genus and ASV-level abundance tables included differential abundance testing, meta-analysis, PERMANOVA, and Random Forest classification to characterize consistent microbial signatures across disease stages.

### Community structure divergence intensifies across diabetes progression

To identify robust microbial biomarkers of diabetes progression and complications, we integrated multiple analytical approaches including community-level comparisons and meta-analysis across independent cohorts. Sample sizes and contributing studies varied across analytical layers due to differences in data resolution and preprocessing requirements. PERMANOVA and community structure analyses incorporated 3–5 studies (n = 156–594 at the genus level), while meta-analysis and differential abundance testing included a broader set of cohorts (n = 198–858) contributing genus-level CLR abundance data, studies included in the analysis, the conditions they represent, and the analyses performed are shown in Table S3.

PERMANOVA analysis of beta diversity revealed significant differences in microbial community composition across disease states, establishing that the gut microbiome undergoes substantial restructuring along the diabetes spectrum. At the genus level, PERMANOVA revealed significant community-wide separation at selected stages of diabetes progression. Significant differences were observed between healthy vs PD (pseudo-F = 3.43, p = 0.015; 3 studies, n = 594) and healthy vs complicated type 2 diabetes (pseudo-F = 3.58, p = 0.001; 5 studies, n = 317) (Figure 1a). In contrast, intermediate transitions showed no significant genus-level effects: PD vs new-onset type 2 diabetes (pseudo-F = 0.947, p = 0.488; 1 study, n = 198), new-onset type 2 diabetes vs type 2 diabetes (pseudo-F = 4.42, p = 0.703; 3 studies, n = 156), and type 2 diabetes vs complicated type 2 diabetes (pseudo-F = 2.32, p = 0.548; 4 studies, n = 345). These non-significant results likely reflect limited sample sizes and inter-study variability. Principal coordinate analysis (PCoA) plots illustrate these genus-level differences (Figure 1b). Healthy vs PD samples show partial but clear separation along the first two axes, consistent with PERMANOVA significance (pseudo-F = 3.43, p = 0.015). Healthy vs complicated type 2 diabetes shows stronger separation (pseudo-F = 3.58, p = 0.001), further confirming robust differences in community structure at the extremes of the disease spectrum.

**Figure 1:**
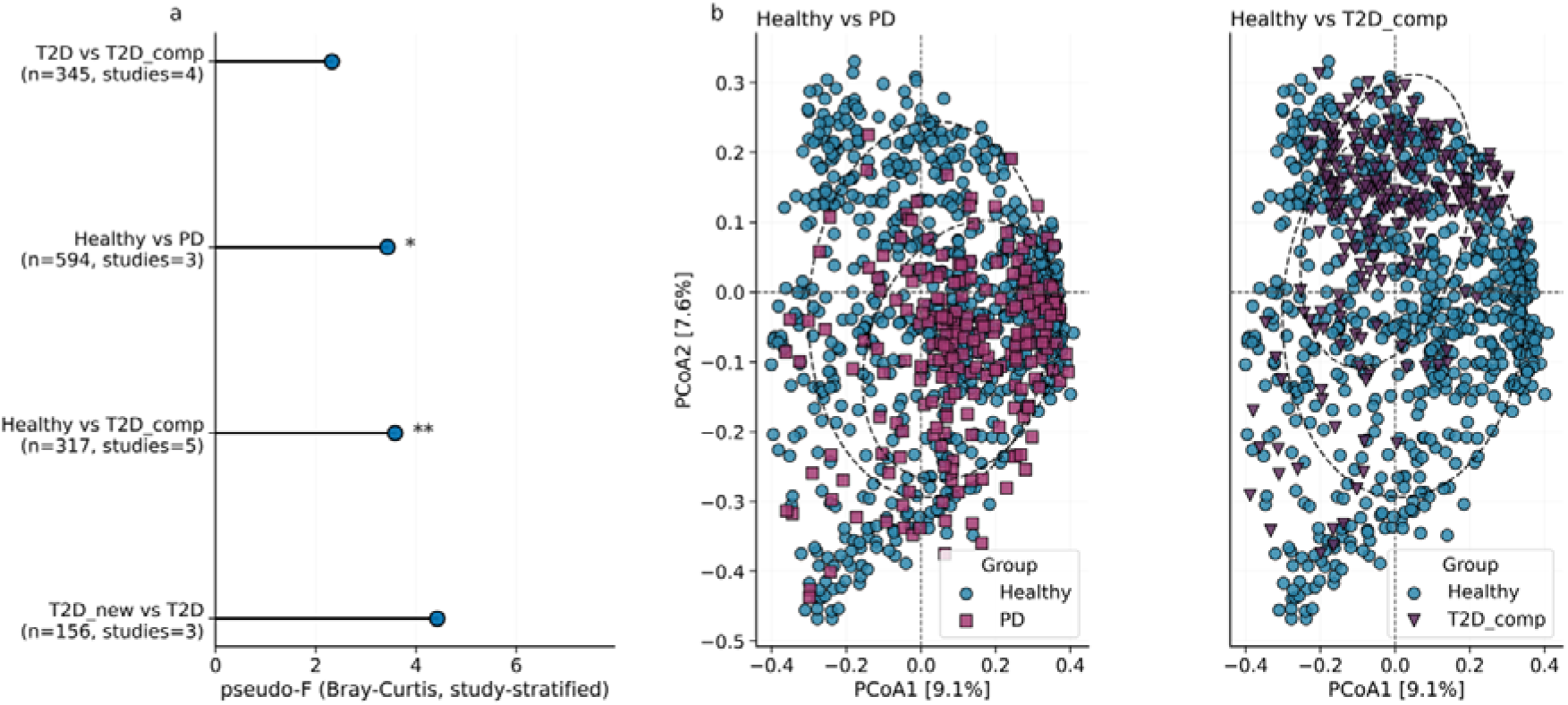
PERMANOVA analysis of gut microbiome composition across diabetes progression. (a) Study-stratified PERMANOVA results (Bray–Curtis dissimilarities) at the genus level. Significant group-level differences were observed for Healthy vs PD (pseudo-F = 3.43, p = 0.015, q = 0.030) and Healthy vs complicated type 2 diabetes (type 2 diabetes_comp) (pseudo-F = 3.58, p = 0.001, q = 0.004), while no significant separation was detected for PD vs new-onset type 2 diabetes (type 2 diabetes_new), type 2 diabetes*_*new vs type 2 diabetes, or type 2 diabetes vs type 2 diabetes_comp. Error bars indicate 95% confidence intervals. (b) Principal coordinates analysis (PCoA) plots illustrating community separation for significant contrasts at the genus level: Healthy vs PD and type 2 diabetes vs type 2 diabetes_comp. Each point represents an individual sample, colored by group, with ellipses indicating 95% confidence intervals.

### Progressive depletion of SCFA-producing genera drives disease-associated restructuring

Having established that community composition differs across disease states, we next identified which specific genera consistently drive these changes. Random-effects meta-analysis of standardised mean differences (Hedges’ g) calculated from CLR-transformed relative abundances identified robust taxonomic biomarkers consistently altered across disease progression. The analysis revealed that SCFA producers including *Faecalibacterium* (*g* = −0.41, *q* = 0.006), *Lachnospira* (*g* = −0.65, *q* = 0.017), *Ruminococcus* (*g* = −0.45, *q* = 0.002), and *Blautia* (*g* = −0.35, *q* = 0.002) were systematically under-represented across disease transitions (Figure 2a). Conversely, opportunistic genera such as *Hungatella* (*g* = 0.44, *q* = 0.002), *Granulicatella* (*g* = 0.38, *q* = 0.001), and *UBA1819* (*g* = 0.40, *q* = 0.006) showed consistent increases in relative abundance from pre-diabetes to complicated type 2 diabetes. Overall, results were highly concordant across studies, exhibiting low-to-moderate heterogeneity (I² = 0–50 %), indicating robust and directionally consistent effects. Additionally, *Escherichia-Shigella* maintained persistently high abundance across all groups, suggesting a background ubiquity rather than a diabetes-specific enrichment.

**Figure 2:**
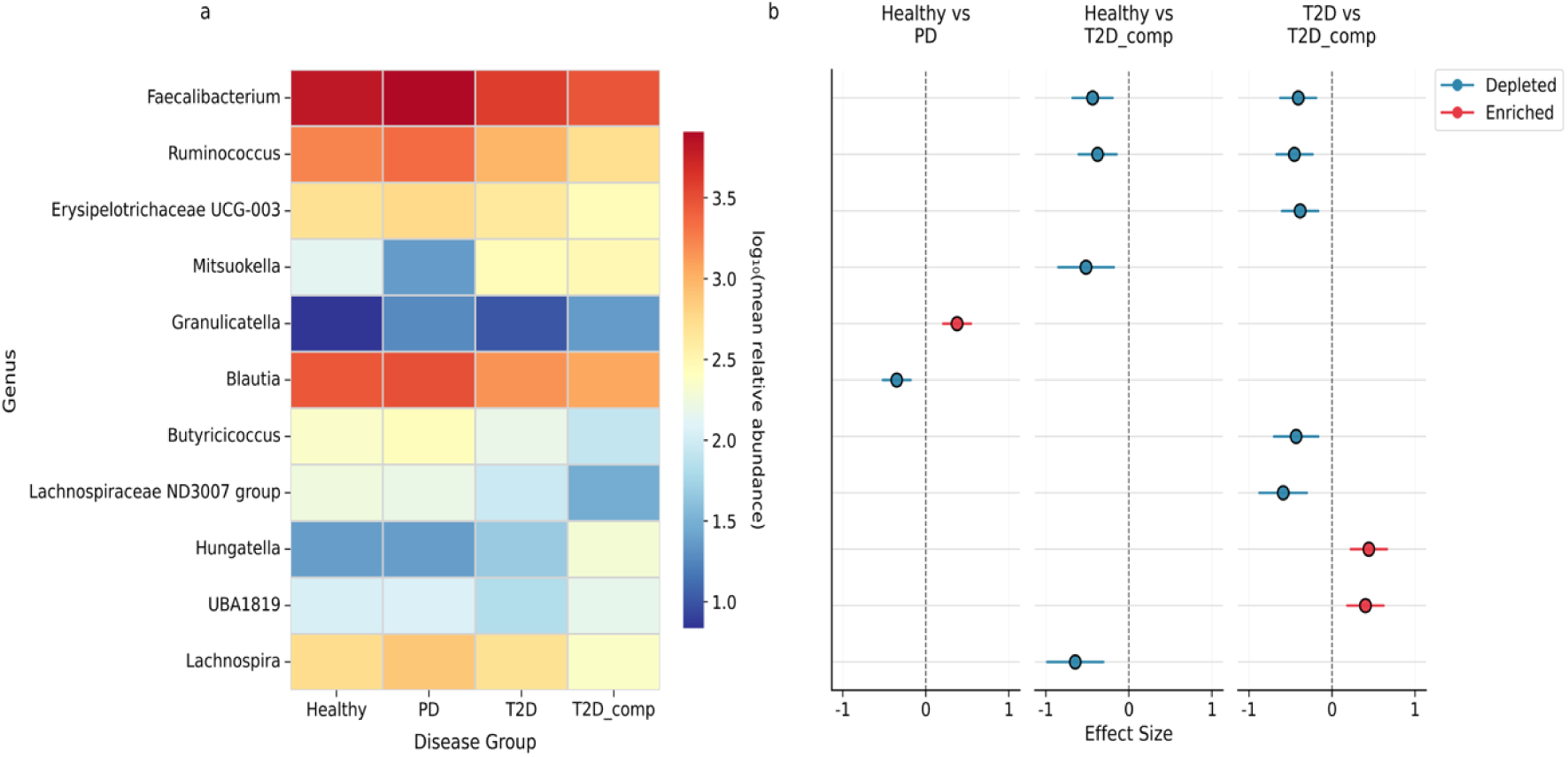
Cross-study meta-analysis of gut microbial genera across diabetes progression. (a) Heatmap showing the log□□-transformed mean relative abundance of meta-significant genera across disease states (Healthy, PD, type 2 diabetes, and type 2 diabetes_comp). Several short-chain-fatty-acid (SCFA)–producing genera, including *Faecalibacterium*, *Ruminococcus*, *Blautia*, *Lachnospira*, and *Lachnospiraceae ND3007 group*, displayed progressive depletion, whereas opportunistic or inflammation-associated taxa such as *Hungatella*, *UBA1819*, *Granulicatella*, and *Erysipelotrichaceae UCG-003* were relatively enriched at later stages. (b) Forest plots summarising pooled meta-analysis effect sizes (Hedges’ g) for the same genera across three key comparisons: Healthy vs PD, Healthy vs type 2 diabetes_comp, and type 2 diabetes vs type 2 diabetes_comp. Blue indicates depletion and red enrichment in disease states. Early alterations were observed for *Granulicatella* (enriched) and *Blautia* (depleted) in prediabetes, followed by pronounced restructuring in advanced diabetes characterised by loss of *Ruminococcus* and *Lachnospiraceae ND3007 group* and enrichment of *Hungatella*.

To ensure that the observed genus-level shifts were consistent across analytical frameworks, we performed complementary ANCOM analyses. ANCOM analysis further supported these patterns, confirming systematic restructuring of the gut microbiota across disease states (Figure S1a).

### Stage-specific microbial transitions reveal early subtle shifts and late marked restructuring

Having identified the overall patterns of SCFA-producer depletion and opportunist enrichment, we next examined the timing and magnitude of these changes at each specific disease transition. In Healthy vs PD (3 studies, n = 594), meta-analysis identified subtle but significant early warning signals with increased relative abundance of Granulicatella (g = 0.376, q = 0.0013, I² = 0%) and significantly reduced relative abundance of Blautia (g = –0.351, q = 0.0025, I² = 0%) and [Eubacterium] hallii group (g = –0.365, q = 0.011, I² = 20%) (Figure 2b). Marked microbial restructuring emerged at advanced disease stages. In type 2 diabetes vs complicated type 2 diabetes (4 studies, n = 345), meta-analysis revealed significantly reduced relative abundance of Lachnospiraceae ND3007 group (g = – 0.588, q = 0.0024, I² = 38%) and Ruminococcus (g = –0.453, q = 0.0024, I² = 0%), alongside increased relative abundance of Hungatella (g = 0.444, q = 0.0024, I² = 0%) and the core biomarker [Clostridium] innocuum group (g = 0.535, q = 0.050, I² = 63%). ANCOM analysis (4 studies, n = 345) identified 22 of 181 tested genera reaching significance, including overrepresentation of the [Clostridium] innocuum group and underrepresentation of Lachnospiraceae ND3007 group and Ruminococcus (W_prop ≥ 0.7) (Figure S1b).

In Healthy vs complicated type 2 diabetes (5 studies, n = 858), the three core biomarkers showed the expected patterns, with meta-analysis confirming significantly reduced relative abundance of Lachnospira (g = –0.647, q = 0.017, I² = 50%) and Faecalibacterium (g = – 0.439, q = 0.017, I² = 8%), alongside increased relative abundance of [Clostridium] innocuum group (g = 0.353, q = 0.049, I² = 0%) (Figure 2b).

Notably, Faecalibacterium, Lachnospira, and [Clostridium] innocuum group emerged as the most robust biomarkers, consistently identified across both analytical frameworks— meta-analysis and ANCOM.

### Microbiome signatures enable robust disease stage discrimination

Blocked Wilcoxon tests revealed stage-specific microbial shifts that were consistent with the patterns identified by meta-analysis and ANCOM discussed above, confirming the robustness of the findings across analytical frameworks.

In the early transition (Healthy vs PD; 3 cohorts, *n* = 594), seven of 63 genera reached significance, including underrepresentation of *[Eubacterium] hallii* group (*q* = 0.02) and *Blautia* (*q* = 0.03), and overrepresentation of *Veillonella* (*q* = 0.03) (Figure 3a). At advanced disease stages (type 2 diabetes vs complicated type 2 diabetes; 3 cohorts, *n* = 211), blocked Wilcoxon tests identified overrepresentation of *Hungatella* (*q* = 0.002) and *UBA1819* (*q* = 0.006), and underrepresentation of *Lachnospiraceae ND3007 group* (*q* = 0.002). Across all comparisons, short-chain-fatty-acid (SCFA)–producing genera including *Faecalibacterium*, *Lachnospira*, *Ruminococcus*, *Blautia*, *Butyricicoccus*, *Mitsuokella*, *Roseburia*, *Dorea*, *Ruminococcus torques group*, and *Clostridia UCG-014* showed consistent declines, whereas opportunistic taxa such as *Granulicatella*, *Hungatella*, and *UBA1819* were enriched, particularly at advanced stages. The test (5 cohorts, *n* = 858) further confirmed underrepresentation of *Lachnospira* (*q* = 0.017) and overrepresentation of *Hungatella* (*q* = 0.002).

**Figure 3:**
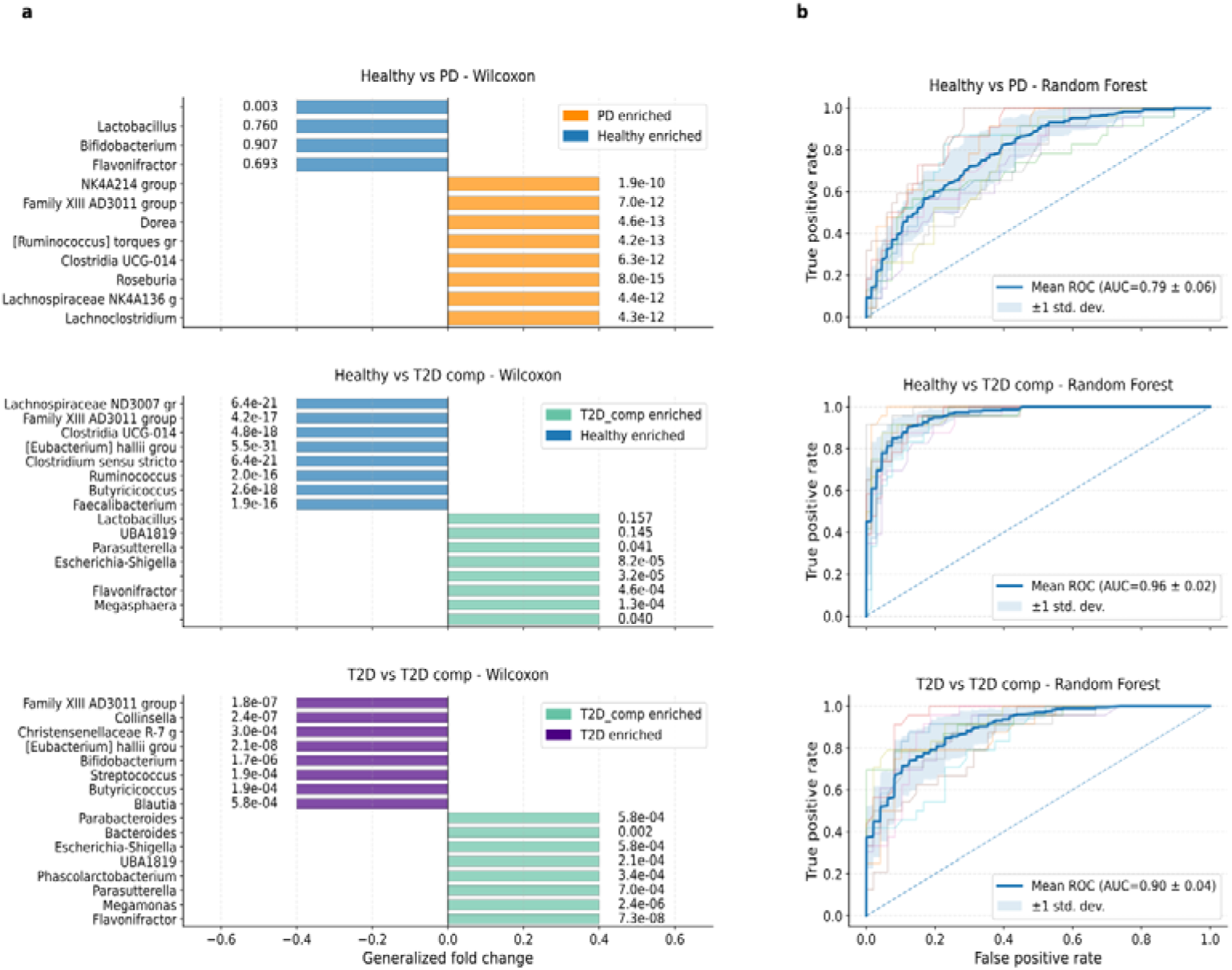
Random Forest validation of Wu-significant genera across disease stages. (a) Differential abundance results from blocked Wilcoxon tests with generalized fold change (gFC) estimates for top genera in each disease comparison (Healthy vs PD, Healthy vs type 2 diabetes_comp, type 2 diabetes vs type 2 diabetes_comp). Bars indicate direction of enrichment (color-coded by group). (b) Random Forest classifier performance for the same comparisons. Receiver operating characteristic (ROC) curves are shown for models trained on Wu-significant genera, with mean area under the curve (AUC) ± standard deviation (blue line ± shaded region). Classifier performance was moderate for Healthy vs PD (AUC = 0.79) but strong for Healthy vs type 2 diabetes_comp (AUC = 0.96) and type 2 diabetes vs type 2 diabetes_comp (AUC = 0.90), confirming robust discriminatory power of taxa such as *Faecalibacterium*, *Lachnospira*, *UBA1819*, *Hungatella*, and *Clostridium innocuum group*.

Genera that were significantly different in blocked Wilcoxon tests (*FDR < 0.05*) were subsequently used as input features for Random Forest classification to assess their discriminatory power across disease stages. Using these Wilcoxon-significant genera, Random Forest models demonstrated strong performance. The Healthy vs complicated type 2 diabetes model achieved an AUC of 0.955, accuracy of 87.6%, specificity of 97.3%, sensitivity of 60.3%, and precision of 88.8% using 76 features (Figure 3b). The type 2 diabetes vs complicated type 2 diabetes model performed robustly (AUC = 0.889, accuracy = 80.9%, specificity = 96.1%, sensitivity = 50.6%, precision = 86.3%) using 22 features. Classification of Healthy vs PD reached moderate performance (AUC = 0.788, accuracy = 76.5%, specificity = 97.3%, sensitivity = 15.4%) using 64 features, consistent with the subtle nature of early-stage microbiome alterations.

### Multiple lines of evidence confirm robustness and reproducibility

To evaluate the robustness and taxonomic resolution of the disease-associated microbiome signatures, we performed complementary sensitivity and ASV-level validation analyses across independent cohorts.

At the genus level, robustness of the Hedges’ g meta-analysis was assessed using two complementary approaches. The per-study comparisons illustrate the direction and magnitude of study-specific Hedges’ g values for each significant genus, enabling direct comparison between individual study effects and the overall pooled genus-level meta-analytic trends (Figure 4a). Most genera showed consistent directionality across studies, with moderate effect sizes (|g| ≈ 0.3–0.6) and low to moderate heterogeneity (I² < 50%), demonstrating the reproducibility of disease-associated shifts across independent cohorts. Quantitative leave-one-out (LOO) sensitivity testing further confirmed the stability of these pooled effects, showing that no single study disproportionately influenced the meta-analytic outcomes (Figure 4b). Across all comparisons, maximum LOO influence values ranged from 0.02 to 0.16, and effect size variations remained below the 0.30 stability threshold, confirming the reliability and reproducibility of the genus-level associations across cohorts.

**Figure 4.**
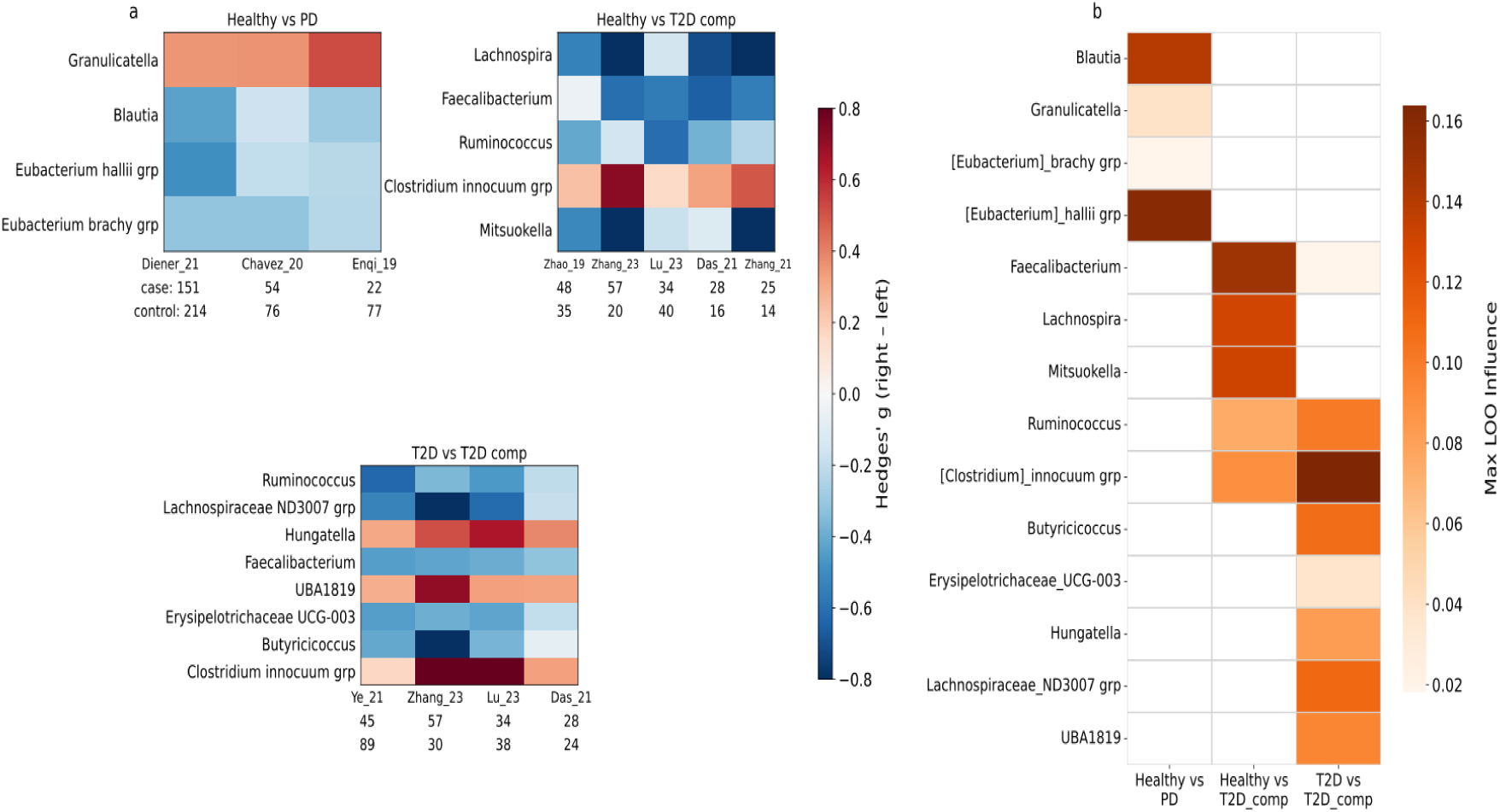
Robustness and sensitivity of genus-level meta-analysis of disease-associated microbiome signatures. *(a)* Study-specific and pooled genus-level effect sizes (Hedges’ *g*) for each significant comparison: *Healthy vs PD*, *Healthy vs type 2 diabetes_comp*, and *type 2 diabetes vs type 2 diabetes_comp*. Each cell represents the standardised mean difference for a genus in a given study, with red indicating enrichment in disease and blue indicating enrichment in controls. White overlays mark genera not meeting the adjusted significance threshold (*q* > 0.05). Case and control sample sizes are shown below each heatmap. Consistent directionality across cohorts with moderate effect sizes (|g| ≈ 0.3–0.6) and low-to-moderate heterogeneity (I² < 50%) indicate robust, reproducible microbial associations. (b) Leave-one-out (LOO) sensitivity analysis of the same genera across comparisons. The heatmap shows maximum influence values quantifying how strongly any individual study affects the pooled random-effects estimate. Darker orange denotes greater influence. Across all genera and comparisons, LOO influence values ranged from 0.02–0.16, well below the 0.30 stability threshold, confirming the robustness and reproducibility of the genus-level meta-analytic results.

At higher taxonomic resolution, amplicon sequence variant (ASV)–level PERMANOVA detected additional significant compositional shifts, capturing differences not apparent at the genus level. Significant separation was observed in four of five comparisons, including type 2 diabetes vs complicated type 2 diabetes (pseudo-F = 2.55, p = 0.027) and Healthy vs complicated type 2 diabetes (pseudo-F = 2.42, p = 0.001) (Table S4). ASV-level meta-analysis further validated these genus-level findings, revealing concordant effect directions for the core biomarker genera across all significant comparisons. In type 2 diabetes vs complicated type 2 diabetes, ASVs belonging to *Faecalibacterium* (q = 0.006–0.049), *Ruminococcus* (q = 0.002–0.030), and *Hungatella* (q = 0.002) matched genus-level trends (Table S5). In Healthy vs complicated type 2 diabetes, multiple ASVs from *Faecalibacterium* (q = 0.005–0.009) and *Lachnospira* (q < 0.05) confirmed consistent underrepresentation. Complementary blocked Wilcoxon tests at the ASV level further supported these findings, with multiple significant ASVs mirroring genus-level patterns. In type 2 diabetes vs complicated type 2 diabetes, significant ASVs within *Faecalibacterium* (q = 0.049), *Ruminococcus* (q = 0.030), and *Lachnospiraceae ND3007 group* (q = 0.046) were consistently depleted. In Healthy vs complicated type 2 diabetes, several *Faecalibacterium* (q = 0.005–0.009) and *Prevotella* (q = 0.001–0.009) ASVs exhibited similar underrepresentation. Together, these analyses demonstrate that the core microbial signatures identified through Hedges’ g meta-analysis are robust, reproducible, and biologically coherent, with consistent validation across both taxonomic levels and independent study cohorts.

## Discussion

Our cross-cohort meta-analysis demonstrates that gut microbiome alterations in type 2 diabetes follow stepwise pattern rather than linear trajectory, with pronounced restructuring only at the extremes of disease progression. Significant compositional shifts were evident between healthy individuals and those with PD or diabetes complications, while intermediate stages showed only subtle changes. Overall diversity declined progressively from healthy controls to complicated type 2 diabetes, accompanied by distinct community separation between groups. Differential abundance and meta-analytic comparisons revealed depletion of butyrate-producing genera and enrichment of opportunistic taxa including. Random Forest models further confirmed that microbial profiles accurately discriminated advanced diabetes and complication states, highlighting robust, reproducible microbiome signatures associated with disease progression. This pattern suggests that microbial instability intensified once metabolic control deteriorates, reflecting a threshold-driven rather than gradual dysbiosis along the diabetes continuum.

Across analytical frameworks, a consistent depletion of SCFA producing genera, including Faecalibacterium, Lachnospira, Ruminococcus, and Blautia was observed. Concurrently, there was an enrichment of opportunistic taxa such as Hungatella, Granulicatella, UBA1819, and the [Clostridium] innocuum group. These compositional shifts reflect a loss of microbial functions essential for intestinal barrier integrity, mucosal immunity, and host metabolic regulation. SCFAs, particularly butyrate and propionate, support epithelial tight-junction stability, modulate GLP-1 and PYY secretion, and supress NF-κB-mediated inflammation [35, 36] Their progressive depletion likely weakens mucosal defense, promotes endotoxemia, and contributes to systemic inflammation and insulin resistance [37]. In early transitions, such as prediabetes, these changes remain subtle, limited to minor reductions in *Blautia*, *Faecalibacterium*, and *Bifidobacterium*, alongside slight enrichment of *Ruminococcus* Hungatella, [Clostridium] innocuum suggesting early functional erosion without overt compositional collapse. Once metabolic dysregulation advances, however, widespread depletion of SCFA producers and expansion of inflammatory genera emerge, marking a threshold beyond which microbial imbalance amplifies rather initiates disease pathology. Mechanistically, SCFA depletion likely disrupts several protective pathways simultaneously: Butyrate fuels colonocytes and enhances tight-junction integrity via claudin-1 and ZO-1 expression [38], acetate, propionate, and butyrate regulate hepatic lipid and glucose flux through AMPK-dependent and PPARγ-mediated pathways [38, 39].

The convergence of findings from meta-analysis, ANCOM, blocked Wilcoxon, and Random Forest classification underscores the robustness of the identified microbial patterns. Core genera, Faecalibacterium, Lachnospira, Ruminococcus, Hungatella, and Clostridium] innocuum, remained consistently significant across all methods of datasets, defining a reproducible microbial signature of disease progression. Random Forest models achieved high discriminatory power for advanced stages (AUC = 0.955 for healthy vs complicated type 2 diabetes), but only moderate accuracy for early transitions, reflecting the weak microbiome signal during initial metabolic disturbance. This specificity pattern indicates greater clinical potential for monitoring progression or complications rather than early detection.

The depletion of butyrate-producing taxa aligns with evidence linking SCFA metabolism to glucose homeostasis and insulin sensitivity [35, 40]. *Faecalibacterium prausnitzii,* a major butyrate producer, was consistently depleted across cohorts, confirming its central role in maintaining metabolic equilibrium. Large-scale human studies similarly associate *Roseburia* and *Coprococcus* with improved insulin sensitivity, supporting a causal link between microbial fermentation capacity and host glucose regulation [35, 41]. in contrast, enrichment of *[Clostridium] innocuum* and *Hungatella* corresponds to increased gut permeability and low-grade inflammation, mechanisms implicated diabetic nephropathy and retinopathy [42–45].

Our stage-specific approach extends prior diabetes meta-analysis that typically compared only healthy and diabetic controls. By resolving intermediate and complicated stages, we demonstrate that reproducible microbial signatures emerge only with escalating metabolic dysfunctional this stratified view clarifies why previous studies have reported inconsistent or cohort-specific findings and underscores that dysbiosis severity parallels disease burden.

This study has some limitations that should be acknowledged. l.The use of 16S rRNA profiling limits taxonomic resolution and functional inference [46], however, the reproducibility across 12 independent cohorts, low heterogeneity (I² < 50%), and stable sensitivity analyses support the robustness of these associations. Integrating metagenomics, metabolomics, and gnotobiotic models will be essential to validate mechanisms, determine causality, and assess the therapeutic value of restoring SCFA-producing consortia or targeting pro-inflammatory taxa [47–49]. Second, potential confounding factors such as diet, medication use, and geographic variation were not uniformly reported across studies, which may have influenced microbiome composition. Third, the cross-sectional nature of the included datasets prevents causal inference regarding microbiome changes and diabetes progression. Nonetheless, the integration of 12 independent cohorts, the consistent results obtained across multiple analytical frameworks, and the low inter-study heterogeneity collectively strengthen the robustness and generalizability of these findings. Third, the cross-sectional design precludes causal inference and temporal assessment of microbiome changes along the diabetes continuum. Although the random-effects meta-analytic framework mitigated inter-study bias and strengthened robustness, longitudinal and mechanistic studies are needed to validate causal links and clarify functional implications of the observed microbial alterations.

This study has some limitations that should be acknowledged. First, this review was limited to 16S rRNA sequencing studies to ensure methodological consistency. While this approach enabled standardised data reprocessing, it excluded shotgun metagenomic studies limiting insights into strain-level and mechanistic interactions [46]. Second, potential confounding factors such as diet, medication use, and geographic variation were not consistently reported across studies, which may have influenced microbiome composition [50–52]. Third, the cross-sectional design precludes causal inference and temporal assessment of microbiome dynamics along the diabetes continuum.

Taken together, this work provides a reproducible, cross-cohort reference map of diabetes-associated microbiome shifts, establishing a foundation for future longitudinal, multi-omics, and mechanistic studies to clarify causality and therapeutic potential.

## Methods

To evaluate microbiome alterations along the diabetes continuum, we identified and selected studies according to PRISMA 2020 guidelines, with the screening and inclusion process summarised in Figure 5a, taxa and community structures were compared across five disease states: healthy, PD, new-onset type 2 diabetes (type 2 diabetes_new), established type 2 diabetes (type 2 diabetes), and complicated type 2 diabetes (type 2 diabetes_comp). Pairwise contrasts were constructed to examine both early transitions (Healthy vs PD) and advanced disease stages (type 2 diabetes vs complicated type 2 diabetes). Complication subtypes, included all samples of diabetic retinopathy (DR), proliferative diabetic retinopathy (PDR), and diabetic nephropathy (DN), pooled into type 2 diabetes with complications (type 2 diabetes_comp) group, enabling identification of stage-specific microbial biomarkers, pipeline details are shown in figure 5b.

**Figure 5.**
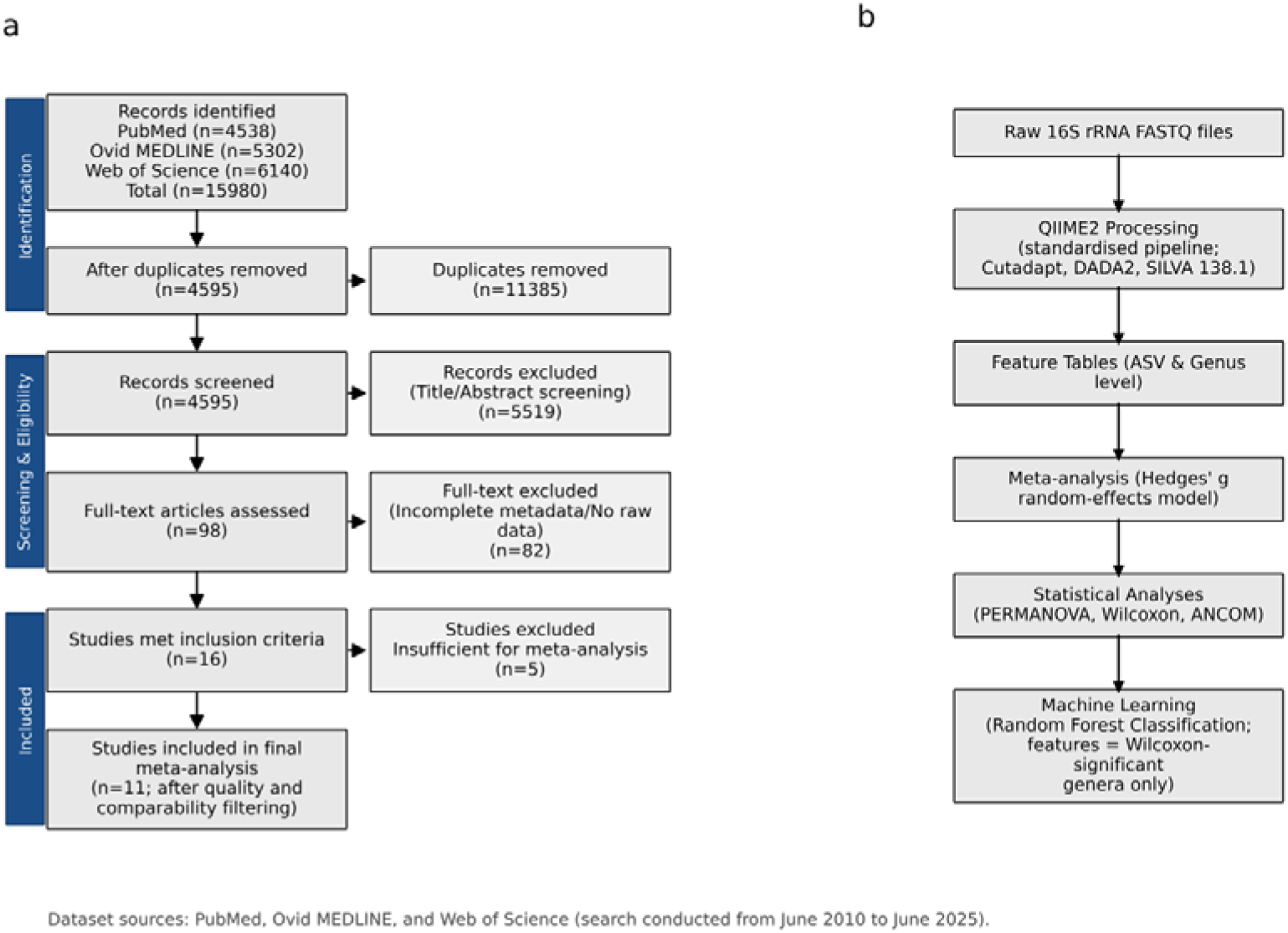
Overview of study selection and analytical workflow. **(a)** PRISMA flow diagram showing the identification, screening, eligibility, and inclusion of studies used in the meta-analysis. Records were retrieved from PubMed, Ovid MEDLINE, and Web of Science between June 2010 and June 2025 **(b)** Overview of the analytical workflow. Raw 16S rRNA sequencing data from twelve independent cohorts were processed through a standardised QIIME2 pipeline (Cutadapt, DADA2, SILVA 138.1) to generate ASV- and genus-level feature tables. A Hedges’ *g* random-effects model was used to integrate study-specific results. Statistical analyses (PERMANOVA, Wilcoxon, and ANCOM) were performed to evaluate community-level and taxon-specific differences across diabetes stages. Finally, Random Forest classification was conducted using Wilcoxon-significant genera as input features to assess their discriminatory power across metabolic phenotypes.

### Study selection and data processing

A systematic search of PubMed, Ovid Medline, and Web of Science databases (search date: June 2025) identified 5,681 unique case-control studies utilising 16S rRNA gene sequencing in type 2 diabetes and prediabetes research (Firgure 5a). Search terms combined keywords and MeSH headings for “gut microbiome,” “microbiota,” “type 2 diabetes,” “prediabetes,” and “insulin resistance”. Both shotgun metagenomic and 16S rRNA sequencing studies were identified in the initial search; however, only 16S rRNA studies were included in the final analysis to ensure a standardised bioinformatic pipeline. To be considered for this work, studies had to follow some required criteria: (i) publicly available raw FASTQ/FASTA sequencing data; (ii) comprehensive metadata with verified case/control status for each sample; (iii) human subjects only; (iv) full-text availability in English; and (v) minimum 15 case patients per study. Two independent reviewers (S.H. and M.E.) screened titles and abstracts in Covidence with automatic duplicate removal and consensus-based conflict resolution. The same reviewers independently extracted data from each included study using Covidence; discrepancies flagged by the software were discussed and resolved by consensus. Detailed search strategies for all databases, registers, and websites, including applied filters and limits, are provided in Table S1. Full-text screening was conducted independently with predetermined eligibility criteria. Raw sequencing data were downloaded from Sequence Read Archive using original publication links. Studies requiring controlled access authorisations or additional ethics approvals were excluded. Primary outcomes were genus-level differences in gut microbial composition across the diabetes continuum. Analyses included PERMANOVA (community-level beta-diversity using Bray–Curtis dissimilarity), ANCOM, and random-effects meta-analysis of standardised mean differences (Hedges’ g) on CLR-transformed abundances. Pairwise differential abundance testing was performed using blocked Wilcoxon tests, and Random Forest classification was applied to genera significant in Wilcoxon analyses. Risk of bias was assessed based on data quality and metadata completeness rather than formal scoring tools. Studies with missing metadata, low read depth, or unbalanced groups were excluded to reduce bias. After rigorous screening, 16 studies were initially selected. Further 4 studies were excluded from the meta-analysis due to quality and comparability issues. These included studies with missing comparison groups (e.g., no healthy samples), low sample sizes (<10 per group), incomplete metadata preventing canonical group mapping, or datasets that yielded no valid genus-level features after filtering. Hedges’ g computation requires at least two comparable groups, and studies failing this criterion were automatically excluded. We got 12 studies ultimately meeting final data quality criteria following bioinformatics processing [20–23, 25–29, 32–34] (Table S2). Data quality was ensured by verifying sequence integrity, metadata completeness, and consistent preprocessing across studies. For differential abundance outcomes, effect sizes were expressed as standardised mean differences (Hedges’ g) on the centered log-ratio (CLR) scale. For community composition (PERMANOVA), pseudo-F statistics and permutation-based p-values were reported. Random Forest classification performance was summarized using AUC, accuracy, precision, sensitivity, specificity, and F1-score. The protocol was registered in the Open Science Framework (OSF; registration https://osf.io/eth7a; embargoed until October 2026) and conducted according to PRISMA guidelines.

### 16S rRNA sequencing data processing

Across all syntheses, the included studies (n = 12) represented diverse geographic regions and sequencing platforms but were uniformly based on human stool 16S rRNA datasets spanning the diabetes continuum as described in Duvallet et al. [15] (Figure 5b). We used QIIME 2 version 2024.2 with region-specific protocols for V3-V4, V4, V4-V5, and V3 amplicons from Illumina and Ion Torrent platforms. Quality control included forward primer trimming using cutadapt plugin (version 2024.2.0, Cutadapt 4.6) with stringent parameters: up to 8 leading ambiguous bases allowed before primer detection, full-length primer overlap required, 7% mismatch tolerance (approximately 1 mismatch per primer), and minimum 50 bp retained read length enforced [53]. DADA2 denoising (plugin version 2024.2.0, R package dada2 1.26.0, R 4.2.2) employed adaptive truncation strategies with standardised lengths of 200 bp (preferred), with stepwise reduction to 150 bp, 120 bp, 110 bp, or 101 bp when higher cutoffs resulted in excessive read loss. Quality filtering used Phred score threshold Q=2 for truncation (e.g. truncating reads when quality is below or equal a score of 2), with maximum expected error thresholds scaled proportionally to read length: 2.0 for 200 bp, 1.5 for 150 bp, 1.2 for 110 bp, and 1.5 for 101 bp reads. Error models were trained on up to 1,000,000 high-quality reads per study to ensure robust statistical modeling. Chimeric sequences were identified and removed using the consensus method with default parameters [54]. Systematic three-step feature filtering was applied per study using feature-table plugin (version 2024.2.2): (1) removal of low-depth samples with <100 total reads; (2) exclusion of rare amplicon sequence variance (ASVs) with <10 total reads across all retained samples; and (3) prevalence filtering to retain only ASVs present in ≥1% of samples within each study, calculated as threshold = ceiling[0.01 × sample size] with minimum threshold of 1 [15]. Representative sequences were filtered post-hoc to match final feature tables. Taxonomic classification employed pre-trained SILVA 138.1 reference classifiers with 99% sequence similarity using feature-classifier plugin (version 2024.2.0) [55]. Region-specific classifiers were separately trained on V3-V4, V4, V4-V5, and V3 amplicon regions and matched to each dataset’s corresponding amplicon target to ensure optimal taxonomic resolution. Classification confidence threshold was set at 0.5, with only sequences assigned to bacterial taxa retained [56].

ASV feature tables were then converted to relative abundances by sample-wise normalisation (dividing by total read depth per sample). Genus-level analyses employed taxonomic collapsing by summing relative abundances within taxonomic rank 6, with unclassified genera excluded from downstream analyses. Both ASV-level and genus-level feature matrices were quality-checked for batch effects and used for all subsequent statistical analyses, differential abundance testing, and machine learning applications.

### Statistical Analysis

All analyses were performed to evaluate community-level and taxon-specific alterations in gut microbiome composition across diabetes progression stages (Healthy, PD, new-onset type 2 diabetes, established type 2 diabetes, and complicated type 2 diabetes). All statistical analyses were conducted in Python v3.8.15 using pandas v1.5.3, numpy v1.24.4 [57], and scipy v1.10.0 [58]. Random-effects meta-analyses were performed with statsmodels v0.14.0 [59], community-level PERMANOVA analyses with scikit-bio v0.5.8, and Random Forest classification with scikit-learn v0.24.1. ANCOM analyses were executed via R v4.2.2. Data visualisation was carried out using matplotlib and seaborn [60, 61]. Multiple testing corrections were applied using the Benjamini–Hochberg false discovery rate (FDR) method [62].

### Community-level analysis

Overall community composition differences were assessed using permutational multivariate analysis of variance (PERMANOVA) on Bray–Curtis dissimilarities [63]. Analyses were conducted at both genus and ASV levels using relative abundance data filtered to retain features present in at least one sample. Only studies with ≥3 samples per group were included per comparison. The pseudo-F statistic was calculated as the ratio of between- to within-group sums of squares, with significance determined by 999 study-stratified permutations. Sample labels were permuted only within studies to preserve cohort structure. P-values were calculated as the proportion of permuted pseudo-F values greater than or equal to the observed statistic [63].

### ANCOM compositional analysis

To validate results under a compositional framework, we applied the original ANCOM (Analysis of Composition of Microbiomes) algorithm [64]. Analyses were run in *Python v3.8.15* and *rpy2 v3.5.11*. Input consisted of genus-level count or relative-abundance tables standardized by metadata. Relative abundances were converted to pseudo-counts (×10,000, rounded) before testing. Taxa present in ≥10% of samples (≥3 observations) were retained. Pairwise comparisons were conducted across all diabetes stages, each with ≥10 samples per group. ANCOM parameters included prevalence cutoff = 10%, minimum library size = 1,000 reads, α = 0.05, and Benjamini–Hochberg FDR correction [62]. A taxon was considered significant if W ≥ 0.7 × (number of taxa − 1). Unclassified taxa were excluded. Results were ranked by W and reported with W_prop (W divided by total taxa).

### Meta-analysis of differential abundance

For each prespecified comparison, genus-level relative abundances were analysed alongside study metadata. Comparisons proceeded only if ≥3 studies met inclusion criteria. To account for compositionality, abundances were centered log-ratio (CLR) transformed after adding a pseudocount of 1 × 10LL [65]. For each taxon and study, standardized mean differences (Hedges’ g) and sampling variances were calculated [57]. Study-level effects were combined using a DerSimonian–Laird random-effects model (method-of-moments τ² estimation), yielding pooled effect sizes, standard errors, z-statistics, p-values, and 95% confidence intervals [66]. Heterogeneity was summarized using Q and I² statistics, and multiple testing controlled with Benjamini–Hochberg FDR [62]. The same procedure was repeated at ASV resolution to improve taxonomic granularity. Leave-one-out (LOO) sensitivity analysis was applied to all taxa with q < 0.10 to evaluate robustness. Results were considered stable if (i) effect direction was consistent across iterations, (ii) pooled effect size variation was <0.30, and (iii) significance (p < 0.05) persisted across all iterations [67].

### Taxon-level differential abundance analysis

Taxon-level differences were tested using a two-sided blocked Wilcoxon rank-sum test, following Wu et al. [17]. Study was treated as a blocking factor to control for inter-cohort variability. Studies were included in a comparison only if both groups had ≥5 samples and at least three eligible studies were available. ASVs were retained if present in ≥20% of pooled samples and detectable in ≥3 studies (any non-zero abundance) [17].

Within each study, relative abundances were ranked using average ranks for ties. The test statistic (T) was calculated as the sum of centered rank sums across studies, defined as T = Σ [R_sum,right − n_right × (N_study + 1)/2]. Significance was assessed using a conditional null distribution generated from 2,000 within-study permutations that preserved group sizes. Two-sided permutation p-values were calculated as p = (count[|T_perm| ≥ |T_obs|] + 1) / (N_perm + 1). Effect sizes were calculated as the mean of per-study median differences.

### Machine Learning Validation

Random Forest (RF) models were trained on genus and ASV level relative abundance profiles merged with metadata to assess discriminatory power of taxa identified by blocked Wilcoxon tests, following Wu et al. [17]. For each comparison, only Wilcoxon-significant genera were used as features, together with available alpha diversity indices (Shannon, Simpson, observed ASVs). Each RF used 501 decision trees, 10% of features per split, and balanced subsample class weights. Performance was evaluated via stratified k-fold cross-validation (maximum 10 folds, determined by smaller group size). Feature selection employed iterative elimination: the least important 10% of features (minimum 1) were removed per iteration until <5 remained or AUC improvement plateaued (ΔAUC ≤ 0.0001 for 5 iterations). Performance metrics included cross-validated AUC, accuracy, precision, sensitivity, specificity, and F1-score. Final models were retrained on full datasets using optimal features, and feature importance values ranked predictors.

### Robustness assessment

To ensure reliability of differential abundance results, two complementary robustness measures were applied. First, amplicon sequence variants (ASVs) were filtered to include only those detected in ≥20% of pooled samples and present in ≥3 independent studies, minimizing the influence of sparsely distributed taxa. Second, a leave-one-out (LOO) sensitivity analysis was performed for all genera with *q* < 0.10, recalculating pooled effect sizes iteratively after excluding each study. Results were considered robust when effect direction remained consistent, pooled Hedges’ *g* variation was < 0.30, and statistical significance (*p* < 0.05) persisted across all iterations.

## Data Availability

All data produced are available online at NCBI

## Data and code availability

All analytic code, summary-level data, and data collection templates used in this study are available from the corresponding author upon reasonable request.

## Use of AI tools

ChatGPT (GPT-5, OpenAI, 2025) was used to assist in code generation and debugging as well as grammar refinement during manuscript preparation. All outputs were reviewed, verified, and edited by the authors to ensure accuracy and integrity.

## Authors contribution

S.H. and T.N.J. conceived the idea. S.H. and M.E. identified and downloaded the datasets. S.H. R.F.D. and S.A. optimised the QIIME pipeline. S.H. optimised downstream analyses, processed the data, interpreted analyses, and wrote the manuscript. T.N.J. and R.F.D. reviewed and provided critical feedback on the manuscript. All authors provided feedback and accepted the manuscript.

## References

1. Hou, K., et al., Microbiota in health and diseases. Signal Transduction and Targeted Therapy, 2022. 7(1): p. 135.

2. Rowland, I., et al., Gut microbiota functions: metabolism of nutrients and other food components. Eur J Nutr, 2018. 57(1): p. 1–24.

3. Loh, J.S., et al., Microbiota–gut–brain axis and its therapeutic applications in neurodegenerative diseases. Signal Transduction and Targeted Therapy, 2024. 9(1): p. 37.

4. Pabst, O., et al., Gut–liver axis: barriers and functional circuits. Nature Reviews Gastroenterology & Hepatology, 2023. 20(7): p. 447–461.

5. Summers, S. and J. Quimby, Insights into the gut-kidney axis and implications for chronic kidney disease management in cats and dogs. The Veterinary Journal, 2024. 306: p. 106181.

6. Bui, T.V.A., et al., The Gut-Heart Axis: Updated Review for The Roles of Microbiome in Cardiovascular Health. Korean Circ J, 2023. 53(8): p. 499–518.

7. Al Bataineh, M.T., et al., Uncovering the relationship between gut microbial dysbiosis, metabolomics, and dietary intake in type 2 diabetes mellitus and in healthy volunteers: a multi-omics analysis. Scientific Reports, 2023. 13(1): p. 17943.

8. Baars, D.P., et al., The central role of the gut microbiota in the pathophysiology and management of type 2 diabetes. Cell Host & Microbe, 2024. 32(8): p. 1280–1300.

9. Zhang, X., et al., Human gut microbiota changes reveal the progression of glucose intolerance. PLoS One, 2013. 8(8): p. e71108.

10. Zhao, L., et al., Gut bacteria selectively promoted by dietary fibers alleviate type 2 diabetes. Science, 2018. 359(6380): p. 1151–1156.

11. Qin, J., et al., A metagenome-wide association study of gut microbiota in type 2 diabetes. Nature, 2012. 490(7418): p. 55–60.

12. Karlsson, F.H., et al., Gut metagenome in European women with normal, impaired and diabetic glucose control. Nature, 2013. 498(7452): p. 99–103.

13. Larsen, N., et al., Gut microbiota in human adults with type 2 diabetes differs from non-diabetic adults. PLoS One, 2010. 5(2): p. e9085.

14. Li, W.Z., et al., Gut microbiota and diabetes: From correlation to causality and mechanism. World J Diabetes, 2020. 11(7): p. 293–308.

15. Duvallet, C., et al., Meta-analysis of gut microbiome studies identifies disease-specific and shared responses. Nature Communications, 2017. 8(1): p. 1784.

16. Wei, Z., G. Chen, and Z.-Z. Tang, Melody: meta-analysis of microbiome association studies for discovering generalizable microbial signatures. Genome Biology, 2025. 26(1): p. 245.

17. Wu, Y., et al., Identification of microbial markers across populations in early detection of colorectal cancer. Nature Communications, 2021. 12(1): p. 3063.

18. Wirbel, J., et al., Meta-analysis of fecal metagenomes reveals global microbial signatures that are specific for colorectal cancer. Nat Med, 2019. 25(4): p. 679–689.

19. Ahmad, A., et al., Analysis of gut microbiota of obese individuals with type 2 diabetes and healthy individuals. PLoS One, 2019. 14(12): p. e0226372.

20. Das, T., et al., Alterations in the gut bacterial microbiome in people with type 2 diabetes mellitus and diabetic retinopathy. Scientific Reports, 2021. 11(1): p. 2738.

21. De, D., et al., Insights of Host Physiological Parameters and Gut Microbiome of Indian Type 2 Diabetic Patients Visualized via Metagenomics and Machine Learning Approaches. Front Microbiol, 2022. 13: p. 914124.

22. Enqi, W., et al., Age-stratified comparative analysis of the differences of gut microbiota associated with blood glucose level. BMC Microbiol, 2019. 19(1): p. 111.

23. Lu, X., J. Ma, and R. Li, Alterations of gut microbiota in biopsy-proven diabetic nephropathy and a long history of diabetes without kidney damage. Sci Rep, 2023. 13(1): p. 12150.

24. Saleem, A., et al., Unique Pakistani gut microbiota highlights population-specific microbiota signatures of type 2 diabetes mellitus. Gut Microbes, 2022. 14(1): p. 2142009.

25. Ye, P., et al., Alterations of the Gut Microbiome and Metabolome in Patients With Proliferative Diabetic Retinopathy. Front Microbiol, 2021. 12: p. 667632.

26. Zhang, L., et al., The Intestinal Microbiota Composition in Early and Late Stages of Diabetic Kidney Disease. Microbiol Spectr, 2023. 11(4): p. e0038223.

27. Zhao, L., et al., Comprehensive relationships between gut microbiome and faecal metabolome in individuals with type 2 diabetes and its complications. Endocrine, 2019. 66(3): p. 526–537.

28. Diener, C., et al., Progressive Shifts in the Gut Microbiome Reflect Prediabetes and Diabetes Development in a Treatment-Naive Mexican Cohort. Front Endocrinol (Lausanne), 2020. 11: p. 602326.

29. Gaike, A.H., et al., The Gut Microbial Diversity of Newly Diagnosed Diabetics but Not of Prediabetics Is Significantly Different from That of Healthy Nondiabetics. mSystems, 2020. 5(2).

30. Kitten, A.K., et al., Gut microbiome differences among Mexican Americans with and without type 2 diabetes mellitus. PLoS One, 2021. 16(5): p. e0251245.

31. Kwan, S.Y., et al., Gut Microbiome Alterations Associated with Diabetes in Mexican Americans in South Texas. mSystems, 2022. 7(3): p. e0003322.

32. Zhang, P., et al., Sex Differences in Fecal Microbiota Correlation With Physiological and Biochemical Indices Associated With End-Stage Renal Disease Caused by Immunoglobulin a Nephropathy or Diabetes. Front Microbiol, 2021. 12: p. 752393.

33. Bhute, S.S., et al., Gut Microbial Diversity Assessment of Indian Type-2-Diabetics Reveals Alterations in Eubacteria, Archaea, and Eukaryotes. Front Microbiol, 2017. 8: p. 214.

34. Chávez-Carbajal, A., et al., Characterization of the Gut Microbiota of Individuals at Different T2D Stages Reveals a Complex Relationship with the Host. Microorganisms, 2020. 8(1).

35. Cui, J., et al., Butyrate-Producing Bacteria and Insulin Homeostasis: The Microbiome and Insulin Longitudinal Evaluation Study (MILES). Diabetes, 2022. 71(11): p. 2438–2446.

36. Wu, D., et al., Cross-Talk Between Gut Microbiota and Adipose Tissues in Obesity and Related Metabolic Diseases. Front Endocrinol (Lausanne), 2022. 13: p. 908868.

37. Yu, W., S. Sun, and Q. Fu, The role of short-chain fatty acid in metabolic syndrome and its complications: focusing on immunity and inflammation. Front Immunol, 2025. 16: p. 1519925.

38. Morrison, D.J. and T. Preston, Formation of short chain fatty acids by the gut microbiota and their impact on human metabolism. Gut Microbes, 2016. 7(3): p. 189–200.

39. Zhang, D., et al., Short-chain fatty acids in diseases. Cell Communication and Signaling, 2023. 21(1): p. 212.

40. Hamari, N., E.E. Blaak, and E.E. Canfora, The impact of butyrate on glycemic control in animals and humans: a comprehensive semi-systemic review. Front Nutr, 2025. 12: p. 1603490.

41. Lv, Q., et al., The role and mechanisms of gut microbiota in diabetic nephropathy, diabetic retinopathy and cardiovascular diseases. Front Microbiol, 2022. 13: p. 977187.

42. Iatcu, C.O., A. Steen, and M. Covasa, Gut Microbiota and Complications of Type-2 Diabetes. Nutrients, 2021. 14(1).

43. Tao, S., et al., Understanding the gut-kidney axis among biopsy-proven diabetic nephropathy, type 2 diabetes mellitus and healthy controls: an analysis of the gut microbiota composition. Acta Diabetol, 2019. 56(5): p. 581–592.

44. Sun, X., et al., Lactiplantibacillus plantarum NKK20 Increases Intestinal Butyrate Production and Inhibits Type 2 Diabetic Kidney Injury through PI3K/Akt Pathway. J Diabetes Res, 2023. 2023: p. 8810106.

45. Huang, Y., et al., Dysbiosis and Implication of the Gut Microbiota in Diabetic Retinopathy. Front Cell Infect Microbiol, 2021. 11: p. 646348.

46. Matchado, M.S., et al., On the limits of 16S rRNA gene-based metagenome prediction and functional profiling. Microb Genom, 2024. 10(2).

47. Byndloss, M., et al., The Gut Microbiota and Diabetes: Research, Translation, and Clinical Applications—2023 Diabetes, Diabetes Care, and Diabetologia Expert Forum. Diabetes Care, 2024. 47(9): p. 1491–1508.

48. Fromentin, S., et al., Microbiome and metabolome features of the cardiometabolic disease spectrum. Nature Medicine, 2022. 28(2): p. 303–314.

49. Crudele, L., et al., Gut microbiota in the pathogenesis and therapeutic approaches of diabetes. eBioMedicine, 2023. 97.

50. Rinninella, E., et al., The role of diet in shaping human gut microbiota. Best Practice & Research Clinical Gastroenterology, 2023. 62-63: p. 101828.

51. Vich Vila, A., et al., Impact of commonly used drugs on the composition and metabolic function of the gut microbiota. Nature Communications, 2020. 11(1): p. 362.

52. Dwiyanto, J., et al., Ethnicity influences the gut microbiota of individuals sharing a geographical location: a cross-sectional study from a middle-income country. Scientific Reports, 2021. 11(1): p. 2618.

53. Callahan, B.J., et al., DADA2: High-resolution sample inference from Illumina amplicon data. Nature Methods, 2016. 13(7): p. 581–583.

54. Estaki, M., et al., QIIME 2 Enables Comprehensive End-to-End Analysis of Diverse Microbiome Data and Comparative Studies with Publicly Available Data. Curr Protoc Bioinformatics, 2020. 70(1): p. e100.

55. Quast, C., et al., The SILVA ribosomal RNA gene database project: improved data processing and web-based tools. Nucleic Acids Research, 2012. 41(D1): p. D590–D596.

56. Yilmaz, P., et al., The SILVA and “All-species Living Tree Project (LTP)” taxonomic frameworks. Nucleic Acids Res, 2014. 42(Database issue): p. D643–8.

57. Hedges, L.V., Distribution Theory for Glass’s Estimator of Effect size and Related Estimators. Journal of Educational Statistics, 1981. 6(2): p. 107–128.

58. Virtanen, P., et al., SciPy 1.0: fundamental algorithms for scientific computing in Python. Nat Methods, 2020. 17(3): p. 261–272.

59. Seabold, S. and J. Perktold, Statsmodels: Econometric and Statistical Modeling with Python. Proceedings of the 9th Python in Science Conference, 2010. 2010.

60. Hunter, J., Matplotlib: A 2D Graphics Environment. Computing in Science & Engineering, 2007. 9: p. 90–95.

61. Waskom, M., seaborn: statistical data visualization. Journal of Open Source Software, 2021. 6: p. 3021.

62. Benjamini, Y. and Y. Hochberg, Controlling the False Discovery Rate: A Practical and Powerful Approach to Multiple Testing. Journal of the Royal Statistical Society: Series B (Methodological), 1995. 57(1): p. 289–300.

63. Anderson, M.J., A new method for non-parametric multivariate analysis of variance. Austral Ecology, 2001. 26(1): p. 32–46.

64. Mandal, S., et al., Analysis of composition of microbiomes: a novel method for studying microbial composition. Microb Ecol Health Dis, 2015. 26: p. 27663.

65. Aitchison, J., The Statistical Analysis of Compositional Data. Journal of the Royal Statistical Society: Series B (Methodological), 1982. 44(2): p. 139–160.

66. DerSimonian, R. and N. Laird, Meta-analysis in clinical trials revisited. Contemp Clin Trials, 2015. 45(Pt A): p. 139–45.

67. Patsopoulos, N.A., E. Evangelou, and J.P. Ioannidis, Sensitivity of between-study heterogeneity in meta-analysis: proposed metrics and empirical evaluation. Int J Epidemiol, 2008. 37(5): p. 1148–57.

