## Supplementary material for "Stage-specific gut microbiome shifts across the Type 2 Diabetes Mellitus spectrum: A systematic review and meta-analysis": Tables S1-S5 and fig S1

**Table S1: Search strategies and results across PubMed, Ovid MEDLINE, and Web of Science for metagenomic and 16S rRNA microbiome studies related to insulin resistance and type 2 diabetes (June 2010 – June 2025).**

| Search Strategy | Database | Search Fields | MeSH Terms Used | Date Searched | Results Retrieved |
| --- | --- | --- | --- | --- | --- |
| Metagenomics + Insulin Resistance | PubMed | Title/Abstract [tiab] | None (text word search only) | 15-Jun-25 | 425 |
|  | Ovid MEDLINE | .mp. (multiple fields) | exp Insulin Resistance/; exp Metabolic Syndrome/ | 15-Jun-25 | 565 |
|  | Web of Science | TS (Topic) | N/A (text word search only) | 15-Jun-25 | 897 |
| 16S rRNA + Insulin Resistance | PubMed | Title/Abstract [tiab] | None (text word search only) | 15-Jun-25 | 627 |
|  | Ovid MEDLINE | .mp. (multiple fields) | exp Insulin Resistance/; exp Metabolic Syndrome/ | 15-Jun-25 | 676 |
|  | Web of Science | TS (Topic) | N/A (text word search only) | 15-Jun-25 | 908 |
| Metagenomics + Type 2 Diabetes | PubMed | Title/Abstract [tiab] | None (text word search only) | 15-Jun-25 | 1,770 |
|  | Ovid MEDLINE | .mp. (multiple fields) | exp Diabetes Mellitus, Type 2/ | 15-Jun-25 | 2,281 |
|  | Web of Science | TS (Topic) | N/A (text word search only) | 15-Jun-25 | 2,469 |
| 16S rRNA + Type 2 Diabetes | PubMed | Title/Abstract [tiab] | None (text word search only) | 16-Jun-25 | 1,689 |
|  | Ovid MEDLINE | .mp. (multiple fields) | exp Diabetes Mellitus, Type 2/ | 16-Jun-25 | 1,780 |
|  | Web of Science | TS (Topic) | N/A (text word search only) | 16-Jun-25 | 1,866 |
| Total (All Strategies) | — | — | — | — | 15,953 |
| After Deduplication (Covidence) | — | — | — | — | 5,681 |

### **Table S2: Data sets collected and processed through standardised pipeline**

| Study | Healthy control | PD | Early-onset T2D | T2D | Complicated T2D | Total | Reference |
| --- | --- | --- | --- | --- | --- | --- | --- |
| **Bhute_2017** | 19 | 0 | 13 | 16 | 0 | 48 | [1] |
| **Chavez_carbajal_2020** | 76 | 54 | 0 | 86 | 0 | 216 | [2] |
| **Das_2021** | 16 | 0 | 0 | 24 | 28 | 68 | [3] |
| **De_2022** | 17 | 0 | 16 | 0 | 0 | 33 | [4] |
| **Diener_2021** | 214 | 151 | 47 | 17 | 0 | 429 | [5] |
| **Enqi_2019** | 77 | 22 | 0 | 33 | 0 | 132 | [6] |
| **Gaikie_2020** | 50 | 0 | 13 | 50 | 0 | 113 | [7] |
| **Lu_2023** | 40 | 0 | 0 | 38 | 34 | 112 | [8] |
| **Ye_2021** | 0 | 0 | 0 | 89 | 45 | 134 | [9] |
| **Zhang_2021** | 14 | 0 | 0 | 0 | 25 | 39 | [10] |
| **Zhang_2023** | 20 | 0 | 0 | 30 | 57 | 107 | [11] |
| **Zhao_2019** | 35 | 0 | 16 | 0 | 48 | 99 | [12] |

**Table S3: Studies included in the analysis, the conditions they represent, and the analyses performed.**

| **Study** | **Healthy vs PD** | **Healthy vs Early_T2D** | **Early_T2D vs T2D** | **T2D_new vs T2D** | **T2D vs T2D_comp** | **Healthy vs T2D_comp** | **Analysis** |
| --- | --- | --- | --- | --- | --- | --- | --- |
| **Bhute_2017** | – | yes | yes | yes | – | – | Meta-analysis, PERMANOVA, Wilcoxon |
| **Chavez_carabjal_2020** | yes | yes | yes | – | – | – | Meta-analysis, PERMANOVA, Wilcoxon |
| **De_2022** | – | yes | – | – | – | – | PERMANOVA |
| **Diener_2021** | yes | yes | yes | yes | – | – | Meta-analysis, PERMANOVA, Wilcoxon |
| **Enqi_2019** | yes | yes | yes | – | – | – | Meta-analysis, PERMANOVA |
| **Gaikie_2020** | – | yes | yes | yes | – | – | Meta-analysis, PERMANOVA |
| **Zhao_2019** | – | yes | – | – | – | yes | Meta-analysis, Wilcoxon |
| **Das_2021** | – | – | – | – | yes | yes | Meta-analysis, PERMANOVA, Wilcoxon, ANCOM |
| **Lu_2023** | – | – | – | – | yes | yes | Meta-analysis, PERMANOVA, Wilcoxon, ANCOM |
| **Ye_2021** | – | – | – | – | yes | – | Meta-analysis, PERMANOVA, ANCOM |
| **Zhang_2021** | – | – | – | – | – | yes | Meta-analysis, Wilcoxon |
| **Zhang_2023** | – | – | – | – | yes | yes | Meta-analysis, PERMANOVA, Wilcoxon, ANCOM |

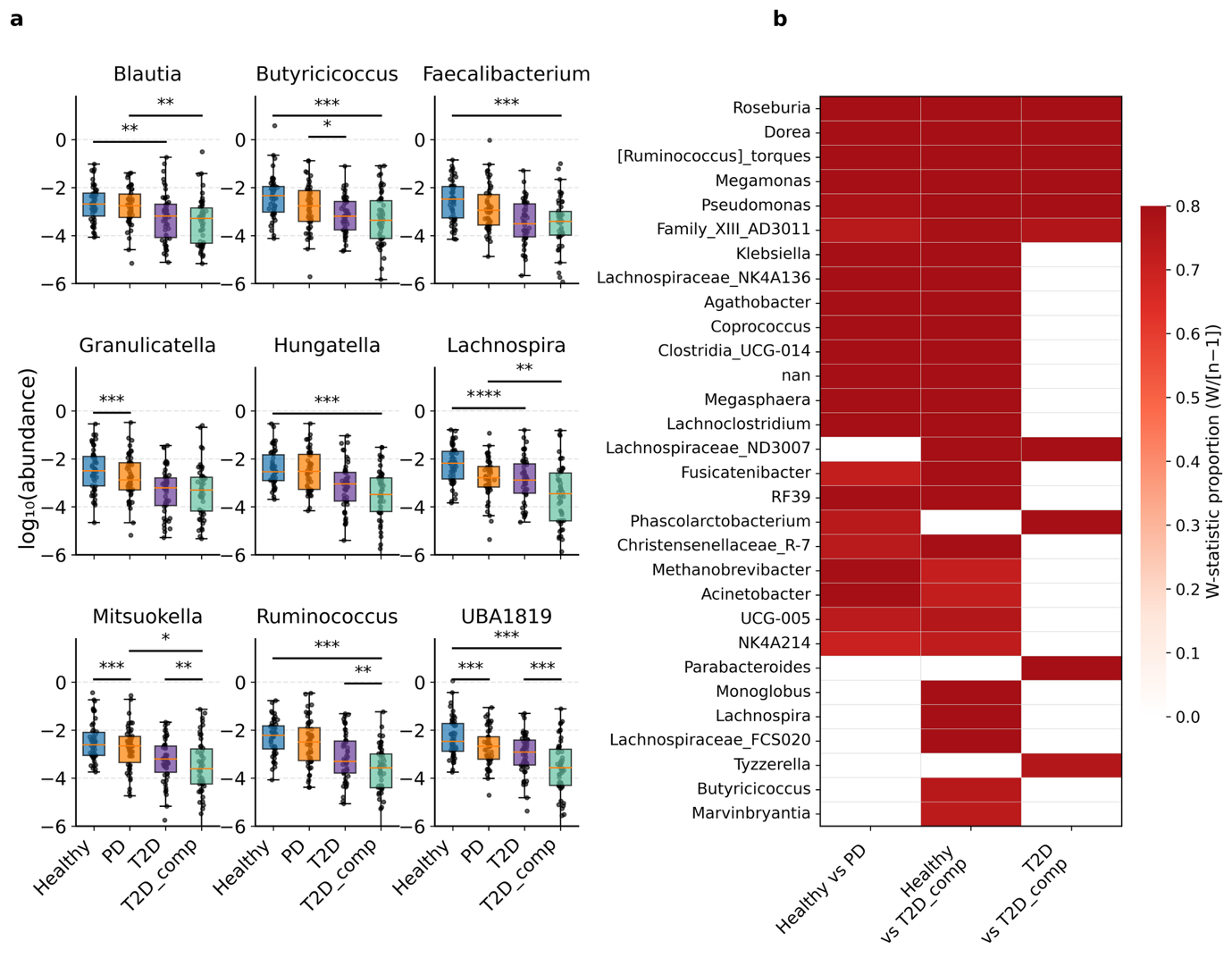

**Figure S1: Robust differential abundance signals identified by ANCOM across diabetes progression.**
(a) Representative genera showing stage-specific abundance patterns across Healthy, Prediabetes (PD), T2D, and T2D with complications (T2D_comp). Short-chain fatty acid (SCFA) producers (Blautia, Butyricicoccus, Faecalibacterium, Lachnospira, Mitsuokella, Ruminococcus) were progressively depleted, while opportunistic taxa (Granulicatella, Hungatella, UBA1819) were enriched in advanced disease. Significance was determined by ANCOM with FDR correction (p < 0.05, *p < 0.01, **p < 0.001).
(b) Heatmap of ANCOM W-statistic proportions (W/[n–1]) across comparisons highlights robust taxa with consistent signals (W_prop ≥ 0.7). Strong effects were observed for taxa including Roseburia, Dorea, Ruminococcus torques group, Clostridia UCG-014, and Lachnospiraceae ND3007 group, reinforcing progressive compositional restructuring with disease severity.

**Table S4: Amplicon sequence variance (ASV) significant from PERMANOVA analysis**

| Comparison | #Studies | n Samples | #Features (ASVs) | pseudo-F | p-value | Interpretation |
| --- | --- | --- | --- | --- | --- | --- |
| Healthy vs PD | 3 | 594 | 28,321 | 2.90 | 0.001 | Significant separation at ASV level |
| PD vs T2D_new | 1 | 198 | 28,321 | 0.855 | 0.716 | Not significant (only 1 study, no cross-study replication) |
| T2D_new vs T2D | 3 | 156 | 28,321 | 6.24 | 0.009 | Significant separation at ASV level |
| T2D vs T2D_comp | 4 | 345 | 28,321 | 2.55 | 0.027 | Significant separation at ASV level |
| Healthy vs T2D_comp | 5 | 317 | 28,321 | 2.42 | 0.001 | Significant separation at ASV level |

**Table S5: ASVs significant from Hedges’ g meta-analysis**

| Comparison | Contributing Studies | N Samples (per group) | Genus / ASV | g | q-value | k | I² | Interpretation | Robustness |
| --- | --- | --- | --- | --- | --- | --- | --- | --- | --- |
| Healthy vs PD | Chavez_carbajal_2020, Diener_2021, Enqi_2019 | Healthy=367; PD=227 | Granulicatella | 0.376 | 0.00129 | 3 | 0% | Higher in PD vs Healthy | yes |
| Healthy vs PD | Chavez_carbajal_2020, Diener_2021, Enqi_2019 | Healthy=367; PD=227 | Blautia | -0.351 | 0.00243 | 3 | 0% | Lower in PD vs Healthy | yes |
| Healthy vs PD | Chavez_carbajal_2020, Diener_2021, Enqi_2019 | Healthy=367; PD=227 | [Eubacterium]_hallii_group | -0.365 | 0.0109 | 3 | 20% | Lower in PD | yes |
| Healthy vs PD | Chavez_carbajal_2020, Diener_2021, Enqi_2019 | Healthy=367; PD=227 | [Eubacterium]_brachy_group | -0.303 | 0.0109 | 3 | 0% | Lower in PD | yes |
| T2D vs T2D_comp | Das_2021, Lu_2023, Ye_2021, Zhang_2023 | T2D=181; T2D_comp=164 | Hungatella | 0.444 | 0.00234 | 4 | 0% | Higher in T2D_comp vs T2D | yes |
| T2D vs T2D_comp | Das_2021, Lu_2023, Ye_2021, Zhang_2023 | T2D=181; T2D_comp=164 | Lachnospiraceae_ND3007_group | -0.588 | 0.00234 | 4 | 38% | Lower in T2D_comp; moderate heterogeneity | no |
| T2D vs T2D_comp | Das_2021, Lu_2023, Ye_2021, Zhang_2023 | T2D=181; T2D_comp=164 | Ruminococcus | -0.453 | 0.00234 | 4 | 0% | Lower in T2D_comp | yes |
| T2D vs T2D_comp | Das_2021, Lu_2023, Ye_2021, Zhang_2023 | T2D=181; T2D_comp=164 | Faecalibacterium | -0.406 | 0.00614 | 4 | 0% | Lower in T2D_comp | yes |
| T2D vs T2D_comp | Das_2021, Lu_2023, Ye_2021, Zhang_2023 | T2D=181; T2D_comp=164 | UBA1819 | 0.402 | 0.00614 | 4 | 0% | Higher in T2D_comp | yes |
| T2D vs T2D_comp | Das_2021, Lu_2023, Ye_2021, Zhang_2023 | T2D=181; T2D_comp=164 | Erysipelotrichaceae_UCG-003 | -0.384 | 0.00918 | 4 | 0% | Lower in T2D_comp | yes |
| T2D vs T2D_comp | Das_2021, Lu_2023, Ye_2021, Zhang_2023 | T2D=181; T2D_comp=164 | Butyricicoccus | -0.432 | 0.0202 | 4 | 30% | Lower in T2D_comp; some heterogeneity | no |
| T2D vs T2D_comp | Das_2021, Lu_2023, Ye_2021, Zhang_2023 | T2D=181; T2D_comp=164 | Unassigned | -0.343 | 0.0251 | 4 | 0% | Lower in T2D_comp (unclassified feature) | yes |
| T2D vs T2D_comp | Das_2021, Lu_2023, Ye_2021, Zhang_2023 | T2D=181; T2D_comp=164 | [Clostridium]_innocuum_group | 0.535 | 0.0497 | 4 | 63% | Higher in T2D_comp; substantial heterogeneity | no |
| Healthy vs T2D_comp | Das_2021, Lu_2023, Zhang_2021, Zhang_2023, Zhao_2019 | Healthy=125; T2D_comp=192 | Lachnospira | -0.647 | 0.0167 | 5 | 50% | Lower in T2D_comp vs Healthy; moderate heterogeneity | no |
| Healthy vs T2D_comp | Das_2021, Lu_2023, Zhang_2021, Zhang_2023, Zhao_2019 | Healthy=125; T2D_comp=192 | Faecalibacterium | -0.439 | 0.0171 | 5 | 8% | Lower in T2D_comp | yes |
| Healthy vs T2D_comp | Das_2021, Lu_2023, Zhang_2021, Zhang_2023, Zhao_2019 | Healthy=125; T2D_comp=192 | Ruminococcus | -0.378 | 0.0390 | 5 | 0% | Lower in T2D_comp | yes |
| Healthy vs T2D_comp | Das_2021, Lu_2023, Zhang_2021, Zhang_2023, Zhao_2019 | Healthy=125; T2D_comp=192 | [Clostridium]_innocuum_group | 0.353 | 0.0483 | 5 | 0% | Higher in T2D_comp | Yes (borderline q) |
| Healthy vs T2D_comp | Das_2021, Lu_2023, Zhang_2021, Zhang_2023, Zhao_2019 | Healthy=125; T2D_comp=192 | Mitsuokella | -0.517 | 0.0483 | 5 | 51% | Lower in T2D_comp; moderate heterogeneity | no |

**References:**

1.	Bhute, S.S., et al., *Gut Microbial Diversity Assessment of Indian Type-2-Diabetics Reveals Alterations in Eubacteria, Archaea, and Eukaryotes.* Front Microbiol, 2017. **8**: p. 214.

2.	Chávez-Carbajal, A., et al., *Characterization of the Gut Microbiota of Individuals at Different T2D Stages Reveals a Complex Relationship with the Host.* Microorganisms, 2020. **8**(1).

3.	Das, T., et al., *Alterations in the gut bacterial microbiome in people with type 2 diabetes mellitus and diabetic retinopathy.* Scientific Reports, 2021. **11**(1): p. 2738.

4.	De, D., et al., *Insights of Host Physiological Parameters and Gut Microbiome of Indian Type 2 Diabetic Patients Visualized via Metagenomics and Machine Learning Approaches.* Front Microbiol, 2022. **13**: p. 914124.

5.	Diener, C., et al., *Progressive Shifts in the Gut Microbiome Reflect Prediabetes and Diabetes Development in a Treatment-Naive Mexican Cohort.* Front Endocrinol (Lausanne), 2020. **11**: p. 602326.

6.	Enqi, W., et al., *Age-stratified comparative analysis of the differences of gut microbiota associated with blood glucose level.* BMC Microbiol, 2019. **19**(1): p. 111.

7.	Gaike, A.H., et al., *The Gut Microbial Diversity of Newly Diagnosed Diabetics but Not of Prediabetics Is Significantly Different from That of Healthy Nondiabetics.* mSystems, 2020. **5**(2).

8.	Lu, X., J. Ma, and R. Li, *Alterations of gut microbiota in biopsy-proven diabetic nephropathy and a long history of diabetes without kidney damage.* Sci Rep, 2023. **13**(1): p. 12150.

9.	Ye, P., et al., *Alterations of the Gut Microbiome and Metabolome in Patients With Proliferative Diabetic Retinopathy.* Front Microbiol, 2021. **12**: p. 667632.

10.	Zhang, P., et al., *Sex Differences in Fecal Microbiota Correlation With Physiological and Biochemical Indices Associated With End-Stage Renal Disease Caused by Immunoglobulin a Nephropathy or Diabetes.* Front Microbiol, 2021. **12**: p. 752393.

11.	Zhang, L., et al., *The Intestinal Microbiota Composition in Early and Late Stages of Diabetic Kidney Disease.* Microbiol Spectr, 2023. **11**(4): p. e0038223.

12.	Zhao, L., et al., *Comprehensive relationships between gut microbiome and faecal metabolome in individuals with type 2 diabetes and its complications.* Endocrine, 2019. **66**(3): p. 526-537.
